# Miami-Dade: A Case Study of Domestic Violence Arrests During the COVID-19 Pandemic

**DOI:** 10.1101/2021.04.20.21255830

**Authors:** Justin Kurland, Alex R. Piquero, Nicole Leeper Piquero

## Abstract

The global health crisis that started early in 2020 has triggered a surge of interest in the effect (if any) of COVID-19 on patterns of domestic violence.^1^ The first systematic review and meta-analysis examining domestic violence during the pandemic revealed quite a lot of diversity in the approaches used to measure potential effects. Drawing on the time series forecasting literature, this brief report contributes to the growing body of evidence around the issue of domestic violence during the pandemic. Arrest data from Miami-Dade County (US) are leveraged along with a robust approach towards model identification, which is used to generate a suitably accurate forecast against which the observed pandemic period domestic violence data can be compared. The pattern uncovered for Miami-Dade County was similar what was found in other U.S. cities that during the pandemic experienced spikes (+95 CI) in the level of domestic violence arrests that were greater than expected. Interestingly these spikes appeared shortly after dips (−95 CI) in observed arrests fell below the expected level.

## Introduction

Since the first quarter of 2020, the coronavirus pandemic has touched the lives of just about every person world-wide. The numbers of infections, hospitalizations, and lives lost are well above any other major pandemic in recent memory. Aside from the public health toll, the pandemic has also shuttered businesses, produced significant unemployment, and disrupted businesses. As well, and for present purposes, the virus and especially its associated lockdown policies have been linked to increases in certain types of crime, in particular homicides and domestic violence.^1,2^

In this study, we further the investigation of how crime, specifically domestic violence, was affected by pandemic lockdowns in one populous but as-of-yet unstudied area, Miami-Dade County, which is home to almost 3 million people, with over two-thirds identifying Hispanic origins. Importantly, our work goes beyond the extant research, especially on domestic violence, in numerous ways. We leverage an approach to forecasting that has hitherto not been leveraged in the extant criminological and criminal justice literature that takes advantage of candidate models, cross-validation and error metrics to identify the most optimal forecast. Further, and importantly, we attempt to describe (quantitatively) extremes in the underlying pattern of domestic violence levels both in terms of both positive and negative spikes. Finally, we evaluate multiple temporal windows, the first that seeks to capture more immediate effects during the early months of the pandemic and then a longer period where changes in the level of stringencies implemented at both the state- and county-level.

In the sections that follow, we provide a brief description of the domestic violence arrest data for Miami-Dade County. We then turn to describing how these data were leveraged to test the more specific hypothesis about whether there were changes in the level in Miami-Dade County that were either greater or less than what would have been expected. The results from the analysis are then presented along with a brief discussion of what this means in relation to the extant literature on domestic violence during the COVID-19 pandemic.

## Methods

### Study Design

This is a time-series study. Using police-recorded domestic violence arrest data for Miami-Dade County between January 2018 and September 2020, we forecasted the daily number of arrests per day for two periods, March 30, 2020 to May 18, 2020 (50 days) and for a longer period between March 30, 2020 and September 14, 2020 (169 days). In both cases, we compared the forecasted number of domestic violence arrests to the observed daily arrests during each of the periods. In this analysis, we selected these two periods because on March 30, 2020, Governor DeSantis of Florida issued a stay-at-home order for South Florida counties including Miami-Dade where over 58% of the state’s cases were concentrated. This order remained in full-effect until May 18, 2020, and thereafter there was a three-phased approach adopted towards an increased, phased in reopening with Miami-Dade County moving into phase two on September 14, 2020.

### Data sources and variables

We obtained the daily domestic violence arrest data from data from the State Attorney General’s Office for the period beginning on January 1, 2018 to September 14, 2020. In accordance with the legal definition as prescribed in the State of Florida *domestic violence*, refers to any assault, aggravated assault, battery, aggravated battery, sexual assault, sexual battery, stalking, aggravated stalking, kidnapping, false imprisonment, or any criminal offense resulting in physical injury or death of one family or household member by another family or household member. Family or household member relates specifically to spouses, former spouses, persons related by blood or marriage, persons who are presently residing together as if a family or who have resided together in the past as if a family, and persons who are parents of a child in common regardless of whether they have been married. With the exception of persons who have a child in common, the family or household members must be currently residing or have in the past resided together in the same single dwelling unit.

In addition to leveraging the arrest data date-based feature engineering was undertaken to account for any seasonality and to improve overall forecast accuracy. For example, a single date, has a day, a day of the week, a day of the year, a week of the year, a week of the month, a month of the year, a month of the series, a year, and a year of the series along with many other combinations and date-based features such as holidays were incorporated into the models.^i^

### Statistical Analysis

The Miami-Dade County domestic violence arrest data was partitioned into three contiguous time-series, twice. The first of the two included a training dataset, for the period between January 1, 2018 and February 8, 2020 (769 days), a 50-day test dataset for the period starting on February 9, 2020 and ending on March 29, 2020, and then our observed period for which we generated the expected forecast (the counterfactual) March 30, 2020 and May 18, 2020 (50 days) but do not use in model building, tuning and evaluation stage of the analytic process. For the second period of interest, the training dataset represented the period starting on January 1, 2018 and ended on October 12, 2019 (650 days), the test period was between October 13, 2019 and ended on March 29, 2020 (169 days) and the forecast period was for the next 169 days from March 30, 2020 to September 14, 2020. Figure 1 below is a visualization of the modeling and forecast process with the dates associated with each respective period. The black points are the observed domestic violence arrests across the entire study period. The red line is a 30-day exponential moving average to help visualize the trend.

**Figure 1:**
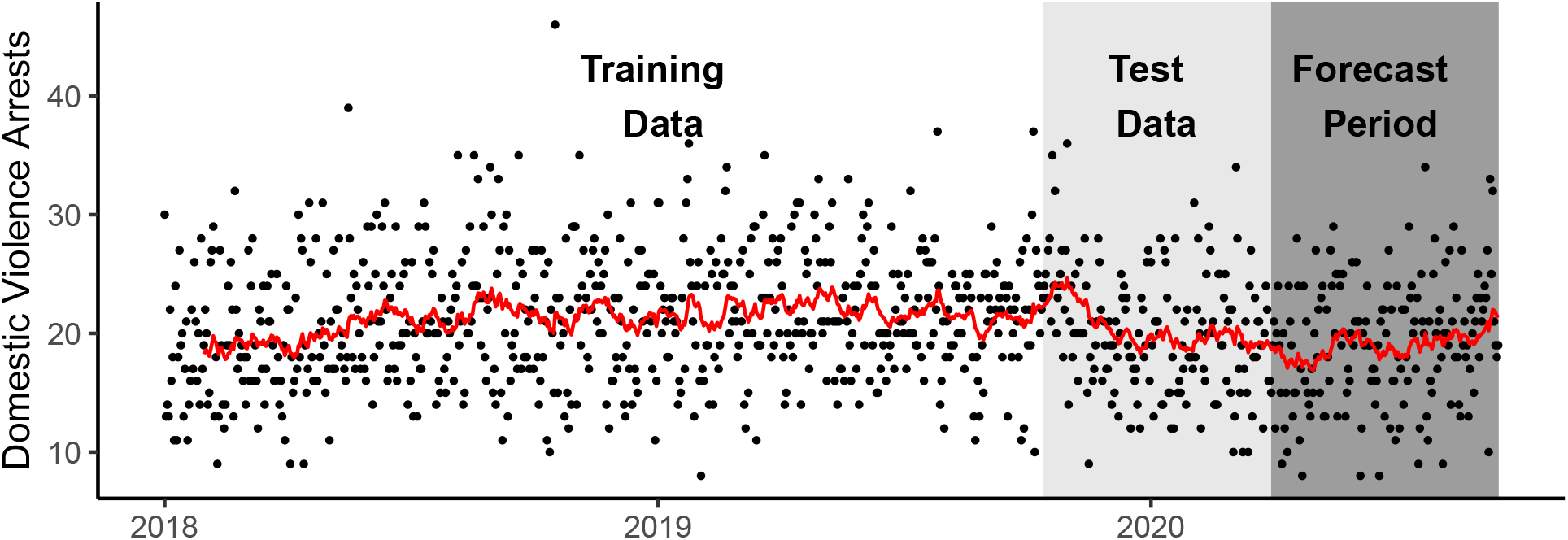
Example of the analytic process that includes the training, test, and forecast period for Miami-Dade County, Florida for the period between January 1, 2018 and September 14, 2020.

In timeseries forecasting it is common for researchers to allocate at least 80% of the original time series to the training dataset and the remainder to the test dataset, in our study we utilized ∼94% for the first period of interest and ∼79% for the second.^3^ The training dataset is used to fit the model, the test dataset provides an unbiased evaluation of a model fit on the training dataset while tuning model hyperparameters, thus providing an unbiased evaluation of a final model fit.^4^ We developed 30 contender models, leveraging both statistical and machine learning approaches such as ARIMA, TBATS, and a feed-forward autoregressive neural network that have been utilized in other related COVID-19 research that has sought to generate forecasts for the test dataset.^5-8^

The point forecasts generated from different forecasting techniques are then compared with the actual data for the test period and the result is used as a proxy measure for accuracy. Numerous error metrics are used here, including Mean Absolute Scaled Error (MASE), Mean Absolute Error (MAE), Mean Absolute Percentage Error (MAPE), Root Mean Squared Error (RMSE), and Symmetric Mean Absolute Percentage Error (SMAPE). These error metrics are ranked for each candidate model and the model with the least error for the test data is selected to produce counterfactuals for the COVID-19 pandemic period, This time, the entire time-series, that is both the training and test datasets were combined and used to build the final model.

## Results

In looking at the arrest data over time you can see there is some daily volatility, but nothing that suggests a level-shift in incidence across this period between January 1, 2019 and September 14, 2020.^ii^ Put differently, the overall trend is relatively stable ranging from a low of roughly 17 to a high of 22 arrests. On average, Miami-Dade County experiences approximately 20 domestic violence related arrests per day, and this varies from 10 to as many as around 30. When looking more closely at the trend it appears that there was a decrease in the level of arrests towards the end of 2019. Importantly, there is a further overall decrease in the trend that coincides with the initial period of the lockdown, but also an increasing trend that is suggestive of a pattern that may be in line with the recent findings from a systematic review/meta-analysis, which found an increase in levels of domestic violence during the lockdown period across numerous locations throughout the U.S. and abroad.^1^

Figure 2 below illustrates the results of the forecast for the first period of interest. The top panel of the firgure shows of the entire timeseries of domestic violence arrests in Miami-Dade (black) and at the end of the series the top performing model forecast (red) along with the associated confidence intervals (grey band). In this case, an ETS (M,A,M) model generated the least error.^iii^ The bottom portion of the plot shows a zoomed in version of the period of interest. Again, the actual arrests for domestic violence can be seen in black, the red line is the forecast, and the grey is the confidence band (±95 CI).

**Figure 2:**
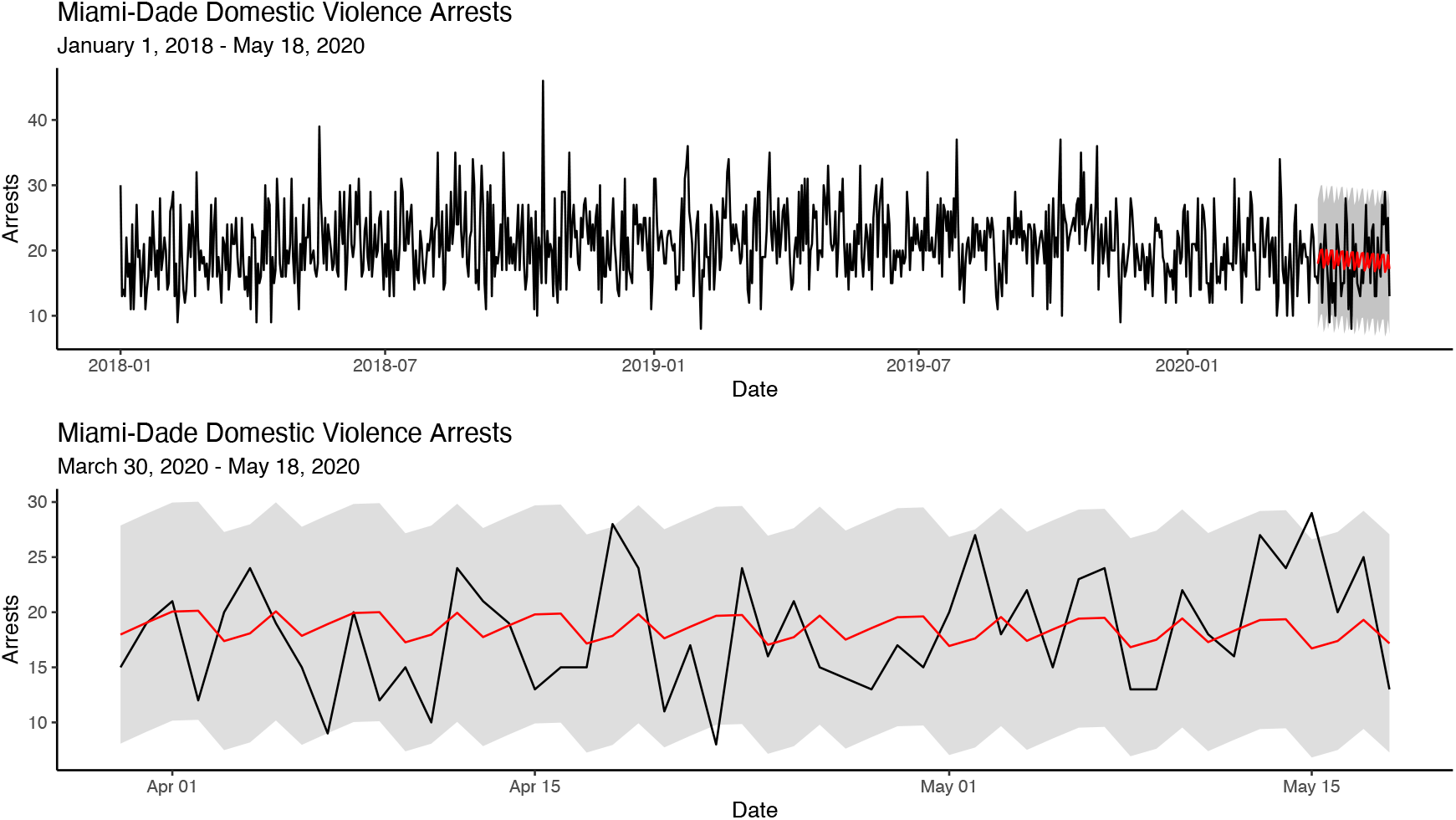
Timeseries for Miami-Dade County for January 1, 2018 to May 18, 2020 with the ETS (M,A,M) forecast for the period between March 30, 2020 and May 18, 2020.

The expectation is that the observed (black line) remains inside the grey band. When the black line crosses either above or below this band these are deviations from the normal process. Put differently, these arrest numbers that suggest that either a significantly greater or lower number of arrests for domestic violence occurred. In this case there were three spikes, a positive spike April 18, followed by negative spike on April 22, and then another positive spike on May 15 where the number of domestic violence arrests exceeded the expected count. The timing of these spikes is in line with other findings during the pandemic in major U.S. cities. That is, a low in arrests and/or calls for service followed by a large spike.

Next, we examine the results, that again, adopted the same approach described above, but ultimately lead to a different model being selected. In this case, a Random Forest (Ranger), model generated the lowest error and can be seen in Figure 3 below. Again, the upper panel of the figure below shows the entire timeseries of domestic violence arrests between January 1, 2018 to September 14, 2020 (black line). The forecast generated for the period of interest between March 30, 2020 and September 14, 2020 (red) can be visualized against the actual count during this period and the confidence band (grey) show the area in which we would expect the number of daily arrests to occur within.

**Figure 3:**
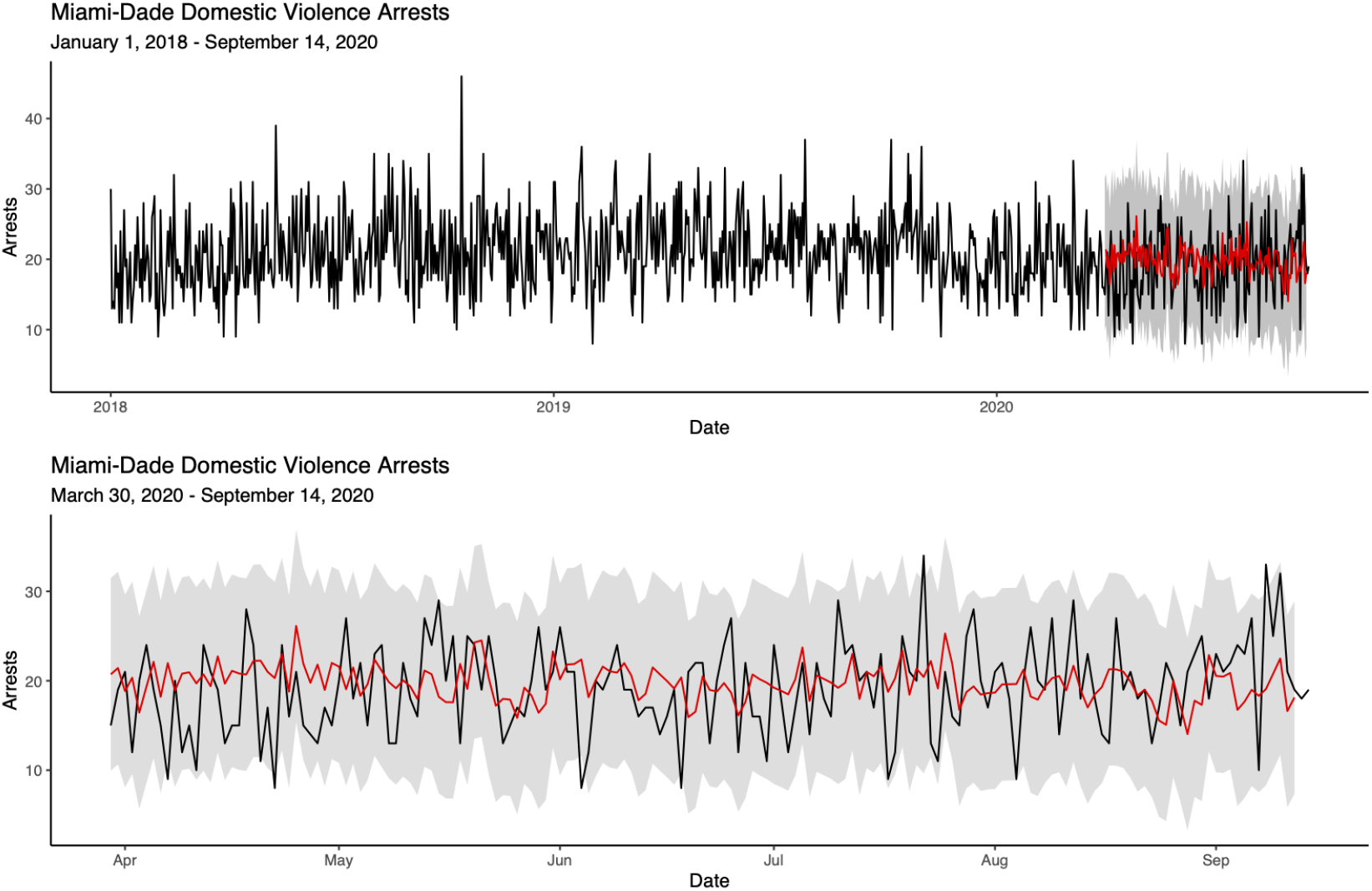
Timeseries for Miami-Dade County for January 1, 2018 to September 14, 2020 with the Random Forest (Ranger) forecast for the period between March 30, 2020 and September 14, 2020.

**Figure 4:**
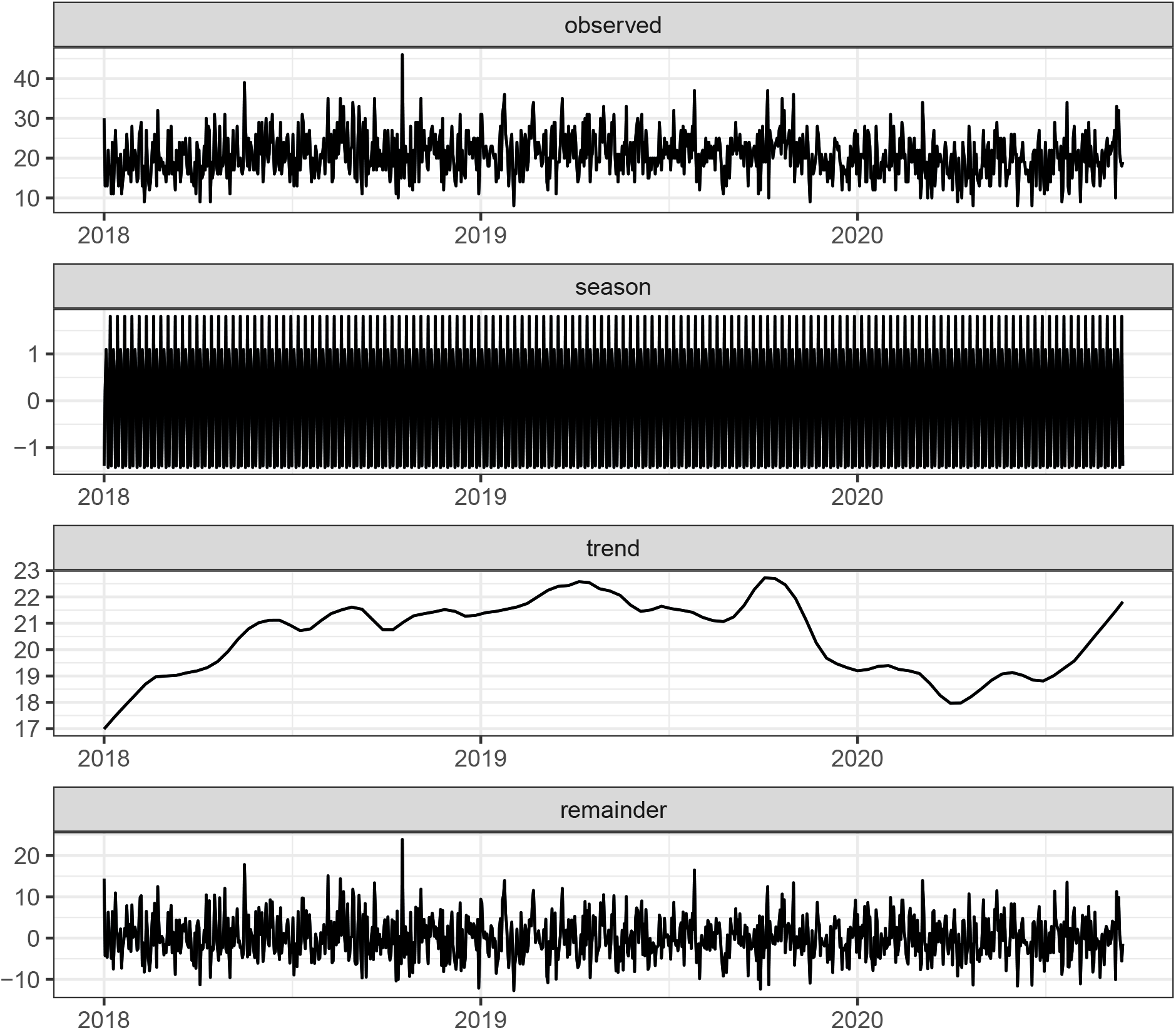
STL decomposition of Domestic Violence Arrests for Miami-Dade County, Florida from January 1, 2018 to September 14, 2020.

**Figure 5:**
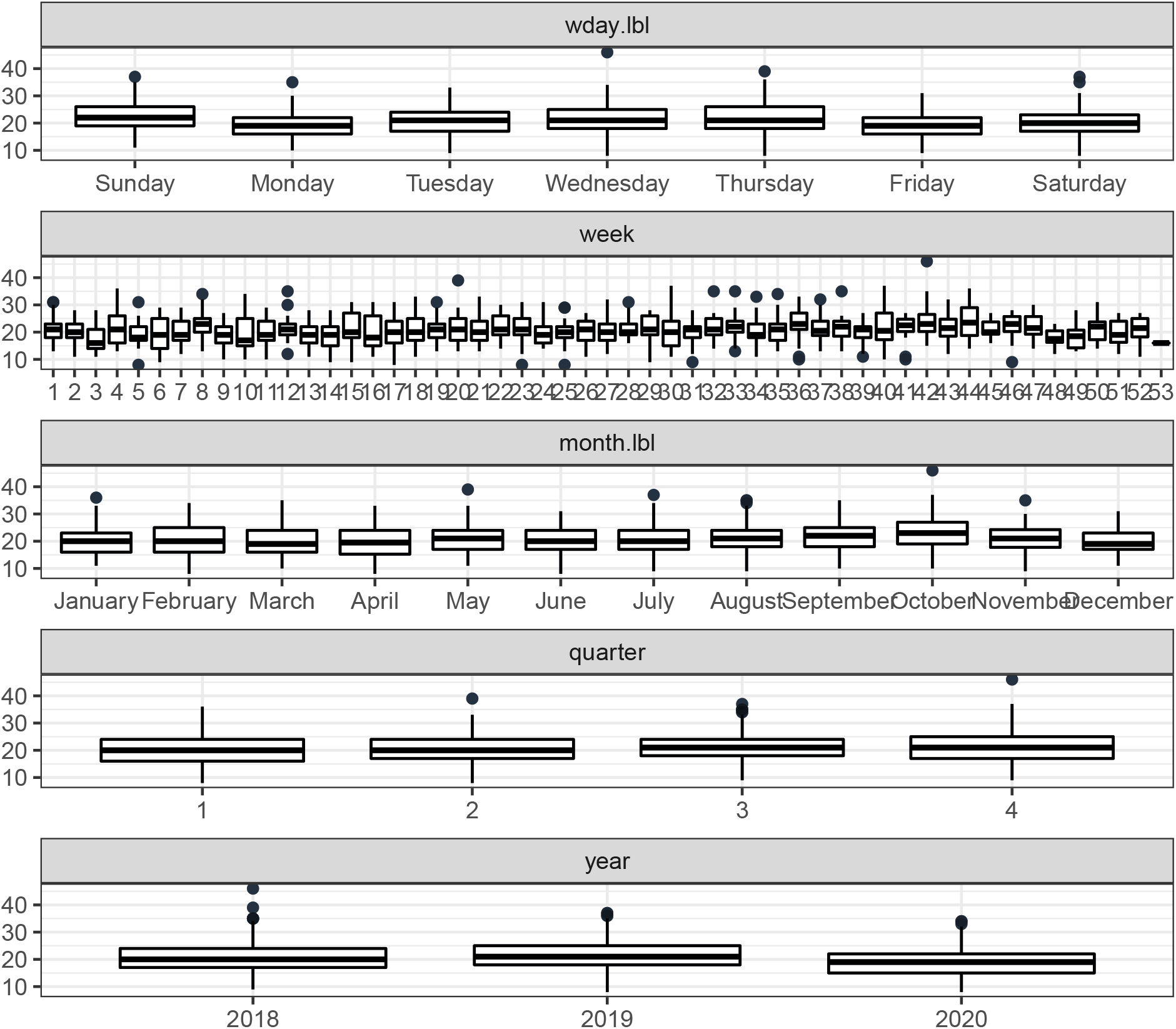
Seasonal Variance of Domestic Violence Arrests for Miami-Dade County, Florida from January 1, 2018 to September 14, 2020 for Day of the Week, Month of the Year, Quarter of the Year, and Year.

The bottom panel shows a zoomed in version of this period of interest. There were five instances where the number of arrests were lower than the confidence bands, these occurred on April 7, 20, and 22, as well as on June 4 and 18, 2020. There were also three instances of spikes exceeding the expected arrest count and these occurred on May 15, July 22, and September 8, 2020. Again, the timing between the spikes is of note. You have a lower than expected number of domestic violence arrests during the early stages of the lockdown in April followed by a peak in May. The pattern then repeats with some lower than expected counts in June, before seeing another spike in arrests shortly thereafter in July.

## Discussion

The coronavirus pandemic has affected just about every aspect of the lives of people around the world. While the coronavirus has affected just about every aspect of the lives of people around th world, our study continues to investigate specific adverse effects of the pandemic, and in particular the lockdowns that were initiated to help stop the spread of the virus on domestic violence. Using Miami-Dade County as our study site, we examined how domestic violence was affected by the pandemic lockdown period.

Adopting a wide variety of methodologies, which heretofore have not been applied to examine this issue, along with a more extensive time period both before and after lockdowns were instituted, the results of our study showed that there were a number of extreme shifts in domestic violence arrests during the lockdown periods that lead to both significantly fewer arrests than what would have been expected, but also and importantly, spikes in the number of arrests. While not causal the lows followed by these highs in arrest numbers in Miami-Dade County provides important guidance around what police, policy-makers, and others involved in helping to reducing injury may look for as indicators in a potential spike in the incidence of arrests during potential future lockdowns.

These results add to the growing knowledge base of how criminal activity, including domestic violence, was altered by the pandemic and public health responses. Our work, however, is not the final word on this issue. For example, we could only examine crime in the form of arrests, and thereby are unable to capture those domestic violence incidents that go undetected and underreported. As well, given the large Hispanic population in Miami-Dade County, more in-depth data should be collected given what is known about the uniqueness of domestic violence within the Hispanic milieu (De Faria, 2021). Lastly, although most of Florida and Miami-Dade County has been open without lockdowns, it will be important to continue to track domestic violence moving forward, especially since people—especially (undocumented) immigrants—continue to feel the pain and anxiety associated with bleak gainful employment prospects and vaccine rollouts.

## Data Availability

Arrest data for Domestic Violence arrests used for this study have been provided by the Attorney General and can be provided to readers upon request.

## What is already known on the subject

- Many cities across the US have experienced spikes in the level of domestic violence during the COVID-19 pandemic.
- School-aged children have also been forced into lockdown creating greater parental responsibility and an added stress during an already volatile situation.

## What this study adds

- Further empirical evidence of spikes in the incidence of domestic violence in Miami-Dade County, Florida.
- A more comprehensive and robust approach to forecasting and assessing injury-related phenomena over time.
- Analysis of a state that remained more open than many others during the ongoing pandemic.

---

**Table 1:** Rank-ordered models by error metrics from cross-validation for the identying the most suitable model for the March 30 and May 18, 2020 forecast.

| Model | MAE | MAPE | MASE | SMAPE | RMSE |
| --- | --- | --- | --- | --- | --- |
| ETS(M,A,M) | 4.140391 | 22.96640 | 0.7643798 | 21.85911 | 5.060820 |
| ETS(A,A,A) | 4.192822 | 23.60763 | 0.7740594 | 22.12095 | 5.074804 |
| ETS(M,N,M) | 4.124940 | 23.90642 | 0.7615275 | 21.73904 | 4.998762 |
| ETS(M,A,N) | 4.163343 | 23.93618 | 0.7686171 | 21.90461 | 5.196125 |
| ETS(M,MD,M) | 4.159794 | 23.94359 | 0.7679620 | 21.92766 | 5.066254 |
| ETS(A,A,N) | 4.170960 | 24.07398 | 0.7700234 | 21.94150 | 5.198306 |
| ETS(M,AD,A) | 4.198104 | 24.18779 | 0.7750345 | 22.11455 | 5.079657 |
| ETS(M,AD,M) | 4.217899 | 24.21664 | 0.7786890 | 22.21713 | 5.087885 |
| ETS(A,AD,A) | 4.212825 | 24.40096 | 0.7777522 | 22.18463 | 5.083011 |
| ETS(M,AD,N) | 4.194260 | 24.44764 | 0.7743250 | 22.05284 | 5.214424 |
| ETS(M,N,A) | 4.193109 | 24.50880 | 0.7741124 | 22.09264 | 5.066435 |
| ETS(A,N,A) | 4.215103 | 24.52606 | 0.7781728 | 22.19272 | 5.097842 |
| ETS(M,M,M) | 4.158979 | 24.65982 | 0.7678116 | 21.89981 | 5.015884 |
| ETS(M,MD,N) | 4.197747 | 24.69073 | 0.7749686 | 22.06408 | 5.219077 |
| ETS(A,AD,N) | 4.198024 | 24.71411 | 0.7750198 | 22.06482 | 5.219804 |
| ETS(A,N,N) | 4.198440 | 24.72484 | 0.7750966 | 22.06662 | 5.220387 |
| ETS(M,N,N) | 4.198740 | 24.74478 | 0.7751520 | 22.06756 | 5.221123 |
| H2O AUTOML - STACKEDENSEMBLE | 4.267674 | 24.82767 | 0.7878783 | 22.43320 | 5.258632 |
| ETS(M,A,A) | 4.223031 | 24.88234 | 0.7796365 | 22.21560 | 5.078026 |
| RANGER | 4.253527 | 25.19878 | 0.7852665 | 22.33991 | 5.233349 |
| RANDOMFOREST | 4.324035 | 25.25904 | 0.7982833 | 22.71320 | 5.295183 |
| ETS(M,M,N) | 4.212789 | 25.33908 | 0.7777456 | 22.12223 | 5.233235 |
| ARIMA(0,1,1)(2,0,0)[7] W/ XGBOOST ERRORS | 4.218204 | 25.39887 | 0.7787453 | 22.14438 | 5.236419 |
| EARTH | 4.420564 | 25.66809 | 0.8161040 | 23.18196 | 5.320920 |
| PROPHET W/ REGRESSORS | 4.335967 | 25.95827 | 0.8004862 | 22.73944 | 5.186241 |
| REGRESSION WITH ARIMA(1,0,2)(2,0,0)[7] ERRORS | 4.378336 | 26.20422 | 0.8083081 | 22.95308 | 5.268169 |
| SEASONAL DECOMP: ETS(A,N,N) | 4.777978 | 26.96728 | 0.8820883 | 24.92753 | 5.768822 |
| NNAR(1,1,10)[7] | 4.865093 | 27.89281 | 0.8981711 | 25.34368 | 5.771463 |
| PROPHET W/ XGBOOST ERRORS | 4.782838 | 28.33972 | 0.8829855 | 24.77462 | 5.919414 |
| LM | 4.614909 | 28.53296 | 0.8519832 | 23.98174 | 5.492105 |
| SEASONAL DECOMP: REGRESSION WITH ARIMA(2,0,1) ERRORS | 5.211406 | 31.20738 | 0.9621057 | 26.62917 | 6.084101 |

**Table 2:** Rank-ordered models by error metrics from cross-validation for the identying the most suitable model for the March 30 and September 14, 2020 forecast.

| Model | MAE | MAPE | MASE | SMAPE | RMSE |
| --- | --- | --- | --- | --- | --- |
| RANGER | 4.174996 | 25.15507 | 0.8194577 | 21.71369 | 5.089200 |
| RANDOMFOREST | 4.204497 | 25.20252 | 0.8252481 | 21.84837 | 5.111900 |
| PROPHET W/ XGBOOST ERRORS | 4.404405 | 25.61556 | 0.8644856 | 22.74739 | 5.486073 |
| H2O AUTOML - STACKEDENSEMBLE | 4.289351 | 25.63554 | 0.8419032 | 22.22721 | 5.187153 |
| ETS(M,A,A) | 4.246980 | 25.83602 | 0.8335866 | 22.00937 | 5.119259 |
| ETS(M,M,N) | 4.269487 | 26.14870 | 0.8380042 | 22.14780 | 5.217254 |
| ETS(A,A,N) | 4.275006 | 26.19337 | 0.8390875 | 22.17289 | 5.222166 |
| ETS(A,N,A) | 4.291521 | 26.22431 | 0.8423290 | 22.20809 | 5.169712 |
| ETS(M,AD,M) | 4.294262 | 26.22505 | 0.8428669 | 22.21790 | 5.182177 |
| ETS(M,A,M) | 4.316476 | 26.24707 | 0.8472270 | 22.33743 | 5.151164 |
| SEASONAL DECOMP: ETS(A,N,N) | 4.339391 | 26.25081 | 0.8517247 | 22.40242 | 5.191982 |
| ETS(M,A,N) | 4.282631 | 26.25544 | 0.8405841 | 22.20752 | 5.228950 |
| ETS(M,N,A) | 4.321745 | 26.50464 | 0.8482613 | 22.34379 | 5.216842 |
| ETS(M,M,M) | 4.331415 | 26.53571 | 0.8501593 | 22.38801 | 5.214725 |
| ETS(M,N,M) | 4.331741 | 26.54602 | 0.8502234 | 22.38166 | 5.230098 |
| ETS(A,N,N) | 4.322464 | 26.57706 | 0.8484025 | 22.38653 | 5.267897 |
| ETS(A,AD,N) | 4.324916 | 26.59615 | 0.8488837 | 22.39754 | 5.270381 |
| NNAR(1,1,10)[7] | 4.387393 | 26.60836 | 0.8611466 | 22.57696 | 5.372621 |
| ETS(M,MD,N) | 4.331958 | 26.65160 | 0.8502659 | 22.42909 | 5.277642 |
| ETS(M,N,N) | 4.333643 | 26.66524 | 0.8505966 | 22.43662 | 5.279437 |
| PROPHET W/ REGRESSORS | 4.582262 | 26.81347 | 0.8993949 | 23.39411 | 5.487979 |
| ETS(M,AD,N) | 4.362142 | 26.88970 | 0.8561903 | 22.56393 | 5.309419 |
| ETS(M,MD,M) | 4.391953 | 27.05533 | 0.8620416 | 22.64707 | 5.307023 |
| ETS(M,AD,A) | 4.421546 | 27.29984 | 0.8678499 | 22.77437 | 5.341981 |
| ETS(A,AD,A) | 4.431098 | 27.38986 | 0.8697249 | 22.81862 | 5.356253 |
| ETS(A,AD,A) | 4.431098 | 27.38986 | 0.8697249 | 22.81862 | 5.356253 |
| ARIMA(0,1,1)(1,0,2)[7] WITH DRIFT W/<br>XGBOOST ERRORS | 4.780374 | 29.85782 | 0.9382798 | 24.38008 | 5.749530 |
| EARTH | 4.936952 | 30.04788 | 0.9690125 | 24.99197 | 6.068195 |
| LM | 4.819589 | 30.27582 | 0.9459769 | 24.50535 | 5.779842 |
| SEASONAL DECOMP: REGRESSION WITH<br>ARIMA(2,0,0) ERRORS | 4.859718 | 30.28553 | 0.9538534 | 24.63105 | 5.822360 |
| REGRESSION WITH ARIMA(1,0,1)(2,0,0)[7]<br>ERRORS | 4.824671 | 30.32667 | 0.9469743 | 24.52909 | 5.784991 |

## Footnotes

i A complete list of all feature engineered variables included in the forecast models is available upon request.

ii Decomposition and seasonality box-plots are provided in the supplementary materials to help readers visualize the trend and seasonality inherent in the Miami-Dade County domestic violence arrest series.

iii The full results of cross-validation for the first and second periods of interest are available in the Supplementary Material for interested readers.

